# Acute Peritoneal Dialysis During the COVID-19 Pandemic at Bellevue Hospital in New York City

**DOI:** 10.1101/2020.08.16.20175992

**Authors:** Nina J. Caplin, Olga Zhdanova, Manish Tandon, Nathan Thompson, Dhwanil Patel, Qandeel Soomro, Fnu Ranjeeta, Leian Joseph, Jennifer Scherer, Shivam Joshi, Betty Dyal, Harminder Chawla, Sitalakshmi Iyer, Doug Bails, Judith Benstein, David S. Goldfarb, Bruce Gelb, Richard Amerling, David M. Charytan

## Abstract

The COVID-19 pandemic created an unprecedented strain on hospitals in New York City. Although practitioners focused on the pulmonary devastation, resources for the provision of dialysis proved to be more constrained. To deal with these shortfalls, NYC Health and Hospitals/Bellevue, NYU Brooklyn, NYU Medical Center and the New York Harbor VA Healthcare System, put together a plan to offset the anticipated increased needs for kidney replacement therapy.

Prior to the pandemic, peritoneal dialysis was not used for acute kidney injury at Bellevue Hospital. We were able to rapidly establish an acute peritoneal dialysis program at Bellevue Hospital for acute kidney injury patients in the intensive care unit. A dedicated surgery team was assembled to work with the nephrologists for bedside placement of the peritoneal dialysis catheters. A multi-disciplinary team was trained by the lead nephrologist to deliver peritoneal dialysis in the intensive care unit. Between April 8, 2020 and May 8, 2020, 39 peritoneal dialysis catheters were placed at Bellevue Hospital. 38 patients were successfully started on peritoneal dialysis. As of June 10, 2020, 16 patients recovered renal function. One end stage kidney disease patient was converted to peritoneal dialysis and was discharged. One catheter was poorly functioning, and the patient was changed to hemodialysis before recovering renal function. There were no episodes of peritonitis and nine incidents of minor leaking, which resolved. Some patients received successful peritoneal dialysis while being ventilated in the prone position.

In summary, despite severe shortages of staff, supplies and dialysis machines during the COVID-19 pandemic, we were able to rapidly implement a *de novo* peritoneal dialysis program which enabled provision of adequate kidney replacement therapy to all admitted patients who needed it. Our experience is a model for the use of acute peritoneal dialysis in crisis situations.

## Introduction

The COVID-19 pandemic created an unprecedented strain on health care systems around the world. Early data from Wuhan, China^1–3^ did not report the high rates of acute kidney injury (AKI) that were subsequently seen in Italy and New York. The dramatic Covid surge in March 2020 in New York City threatened to overwhelm hospital capacity^4,5^ for provision of kidney replacement therapy (KRT). During early March 2020, NYC Health + Hospitals/Bellevue (BH) put together an action plan (Supplementary Table 1) to manage the anticipated increased needs for KRT; acute peritoneal dialysis (PD) was thought to be the best option to rapidly expand the capacity to provide KRT. By April 1, 2020, it became clear that our ability to handle the surge of patients with AKI using our current modalities, intermittent hemodialysis (IHD) and continuous venovenous hemofiltration (CVVH), was insufficient. Many hemodialysis nurses were unavailable due to COVID-related illness resulting in a shortage of trained nurses. The intensive care unit (ICU) nursing staff that performs CVVH was overtaxed because of the expansion of ICU capacity mandated by New York State. Furthermore, CVVH machines were being used at full capacity and CVVH supplies were rationed by the supplier and being rapidly depleted.

In response, we promptly implemented the plan that had been conceived weeks prior to the surge. Under normal circumstances in Bellevue, acute PD had not been utilized for AKI for several decades. According to the International Society of Peritoneal Dialysis (ISPD) guidelines, the use of PD to treat patients with AKI is an acceptable form of treatment.^6^ Given the urgent nature of the circumstances, we felt the necessity to establish acute PD capability as our usual KRT modalities were being overwhelmed by COVID-19 AKI. Herein, we describe the rapid and successful implementation of an acute PD program during the COVID-19 pandemic in a period from mid-March to May 2020.

## ICU Capabilities and Initial Challenges for KRT

Prior to the COVID-19 pandemic, BH had 66 ICU beds, 10 ICU-capable beds in the Emergency Department (ED) and 780 total beds. The inpatient hemodialysis (HD) unit could accommodate 12 inpatients per day, six days per week. Three portable HD machines and four CVVH machines were sufficient to provide bedside dialysis in both the ICU and non-ICU inpatients.

A total of 51 patients were seen by the Nephrology Service between January 20 – February 1, 2020; 26 were ESKD patients who received dialysis in the inpatient unit, 9 were ESKD patients who received bedside dialysis. 11 patients were evaluated for AKI and need for KRT in the ICU; 5 received bedside IHD, 3 received CVVH and 3 did not require CVVH. Typically, fewer than 5 patients per day received bedside KRT between the ICU and regular floors.

In response to the surge, 90 overflow ICU beds were established in ICUs, ED, the endoscopy suite, and retrofitted old wards. At its peak on April 7, 2020, Bellevue had 134 ICU patients and 25 additional critical care patients in an overflow ICU area in the ED. Between March 10 to May 17, 2020, the Nephrology Service evaluated a total of 159 ICU patients with stage 2 or 3 AKI, most requiring KRT. This was not expected and was above the previously reported levels of COVID-19 associated AKI.^4,5^

Additionally, the inpatient HD unit was closed for COVID patients, necessitating bedside HD for all admitted patients as most non-COVID patients were transferred to outside hospitals. During this time, we accommodated 35–40 inpatients requiring KRT per day, 15–20 of whom received bedside hemodialysis treatments (Figure 1).

**Figure 1:**
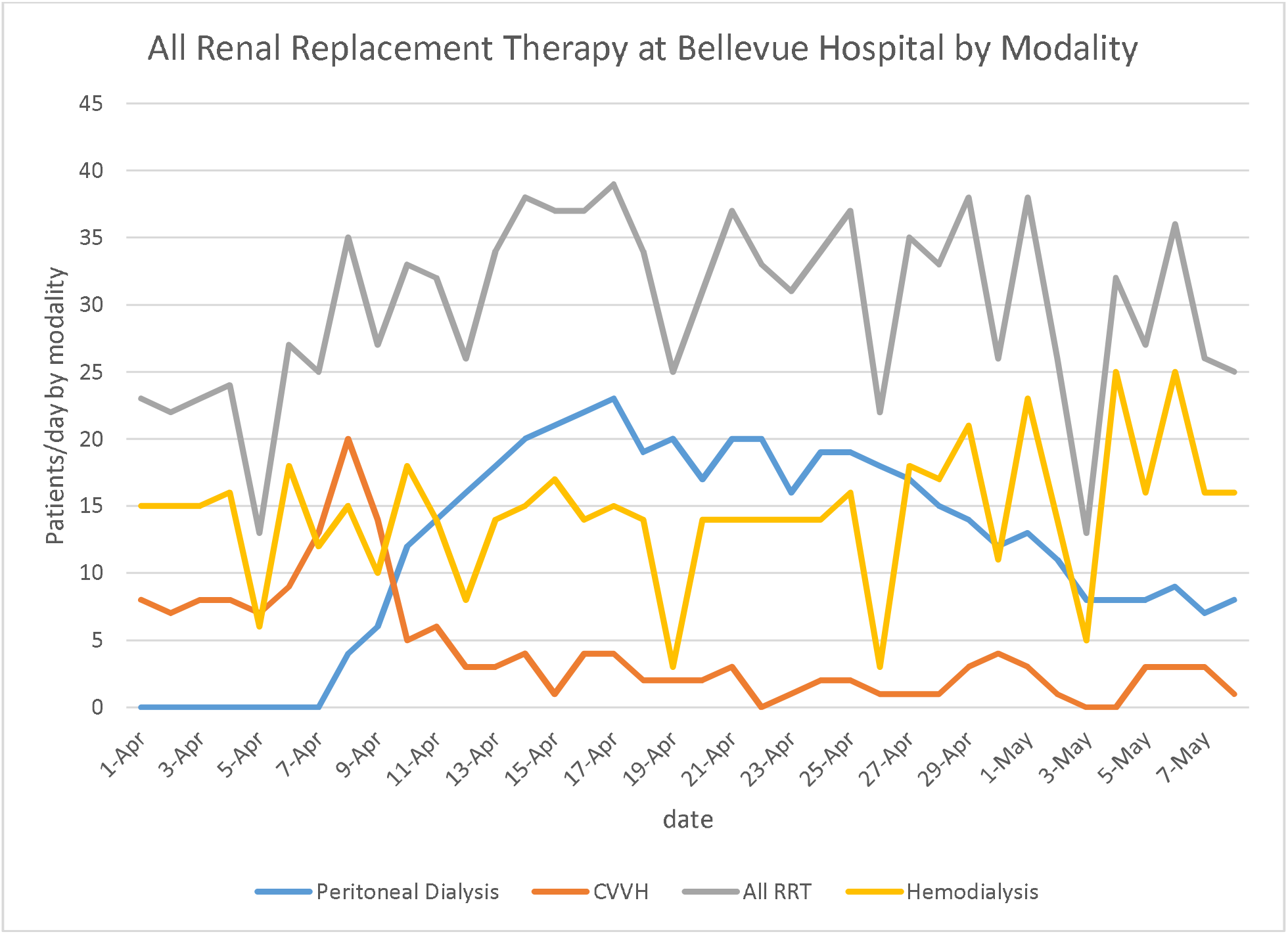
Dialysis modalities between April 1, 2020 and May 8, 2020 at Bellevue Hospital Center; KRT: kidney replacement therapy; CVVH: continuous venovenous hemofiltration.

The number of patients requiring KRT exceeded our baseline capacity for both IHD and CVVH. The challenges for providing IHD were (1) inadequate number of portable HD machines, (2) insufficient staffing due to illness and increased census, and (3) lack of adequate plumbing for water sources and usable drains in newly created ICU areas. Provision of CVVH was also severely limited for the same reasons.

We attempted to expand our KRT capabilities in several ways. Hemodialysis treatments were shortened and the frequency decreased to less than three times per week for selected patients based on their metabolic and volume requirements. Non-COVID stable chronic HD patients were transferred out to affiliated facilities. CVVH was expanded by performing two 10 hour accelerated venovenous hemofiltration (AVVH) treatments per one machine in 24 hours, providing two patients with treatments each day.^7,8^ However, this strategy rapidly depleted CVVH disposable supplies (filters, tubing, dialysate bags, disposable bags) with no prospect to replete due to nationwide rationing imposed by suppliers. COVID19 infected patients also had an increased propensity to clot HD blood lines, as well as IHD and CVVH membranes and circuits, rendering these modalities useless in some patients and contributing to supply shortages. These patients experienced worsened anemia, further driving demand for blood products which were in short supply. We were facing the prospect of rationing dialysis resources.

To address these issues, we created an acute PD program. Initial planning took place in the weeks prior to the surge and took about 2 weeks to implement, with the first PD catheter placed on April 8, 2020. The acute PD program turned out to be instrumental in the BH response to COVID-associated AKI.

## Planning

The use of acute PD to treat AKI requiring KRT was non-existent prior to the COVID-19 pandemic at BH. Consistent with the overall practice trends in the United States, there were very few chronic PD patients as well. However, in anticipation of greater need for KRT during the surge of COVID-19 patients, we put together an acute PD implementation protocol (Supplementary Table 1) in discussion with colleagues internally and from other institutions. The protocol described the roles and responsibilities of the staff, included supply lists, guidance for PD catheter placement, and links to online training resources for performing continuous ambulatory PD (CAPD), automated PD (APD), and percutaneous catheter placement.

Training of staff was a priority since there were few able to perform PD. We had to educate ICU doctors and nurses about acute PD in AKI and the equivalence of PD to other KRT modalities, and how its use would avoid rationing.^9–12^

## Supply Chain Issues

Before the surge, we accrued a list of PD supplies with the assistance of experienced PD nurses. We were able to find vendors who rapidly supplied us with solutions, transfer sets, drain bags and PD catheters. We obtained about 100 catheters of different sizes to ensure sufficient capacity and to reduce the need to restock in the middle of the pandemic. With elective surgery suspended and operating supplies available, surgeons were able to assemble catheter insertion instrument trays.

## Surgical Support

A team of surgeons committed to provide support with insertion and management of PD catheters was an essential part of the plan. The lead surgeon (MT) at BH was responsible for finalizing the details of the insertion technique and acted as a point person for all procedures to

Acute Peritoneal Dialysis for COVID-19 increase efficiency. The team of surgeons was available around the clock 7 days a week, allowing PD catheters to be placed within 12 hours of request by the nephrology team. The catheters were primarily inserted using a limited cut down to the peritoneal membrane through the rectus muscle at bedside in the ICU as all but one patient was intubated and sedated.^13,14^ Laparoscopic technique was not employed because of potential aerosolization of COVID-19 particles.^15^

## Staffing, Staff Training and Initial Experience

Major advantages of PD are its low-tech nature and relative ease for rapid training. This was critical given the constraints of trained nursing staff noted above. The initial PD team consisted of the lead nephrologist (NC), volunteer non-nephrology physicians (including pediatric ophthalmologists and a dermatologist), and ambulatory care nurses. Subsequently, the team included volunteer PD nurses, nurse practitioners, and physician assistants obtained through the Federal Emergency Management Agency (FEMA).

An experienced PD nurse from a private outpatient dialysis unit affiliated with BH and the lead nephrologist made training videos for manual PD and for automated PD. Lessons from the nephrologist and the ‘homemade’ training videos were used to train the new PD team. Online resources from Fresenius and Baxter (see supplement 1) were also utilized for additional detail but were not tailored for our acute PD needs. Team members were familiar with the main aspects of sterile procedures due to their medical background and were able to effectively learn the sterile procedures needed for PD. Overall, 25 people were on the PD team and we were able to provide exchanges 24 hours per day by the end of the first week. Some of the team members from FEMA were PD nurses who also assisted with hands-on training and supervision.

## PD Prescription and Delivery

PD catheters were flushed and used immediately after insertion with low volume exchanges (500 mL). The freshly inserted catheter was flushed 3 times with 500 mL of 1.5% dextrose solution or until clear if bloody. Heparin, 500U/L was added to dialysate to prevent fibrin formation. The initial exchange volume was 500 mL of 1.5% or 2.5% dextrose solution with a dwell time of 2 hours. In the absence of leaks, we increased exchange volume by 250 mL every 2–3 exchanges for the first 6 exchanges then more rapidly until a volume of 2000 mL was reached, usually within the first 36 hours. In the event of leaks, dwell volume was reduced, or exchanges were held for 12 hours. The typical PD prescription was 5–8 exchanges per day, depending on dwell time, over 17 hours. As team members were added, we expanded PD exchanges to 24 hours and were able to achieve higher clearance using manual PD until cyclers were available. The typical exchange volume was 10–16 L per day when manual PD was used; exchange volume increased to 17–20 L per 24-hour period when cyclers were used. Adjustments to these prescriptions were made according to individual patient ultrafiltration and metabolic needs.

In mid-April, we acquired 18 automated cyclers which greatly eased the workload of the PD team and enabled high volume PD for better clearance.^16,17^ Patients who had functioning PD catheters who were in the supine position were subsequently placed on cyclers following our initial manual prescription to ensure the catheter was functioning well.

Patients in the proned position remained on manual exchanges because occasionally flow was obstructed, and was more easily adjusted with manual exchanges. This issue occurred less frequently with more experience and coordination with the proning team. In total, seven patients received PD while being placed in the prone position for 19 hours per day, one of whom recovered kidney function. The prescription was adjusted for these patients with manual exchanges every 1 hour while supine and every 2–3 hours while proned with a maximum 1500 mL dwell while in the prone position. We were able to successfully perform adequate manual PD on patients who were proned with minimal complications by carefully coordinating with proning teams.^16,17^

Patients were moved closer to the door, enabling the cyclers to be placed outside of the room (Figure 2), to minimize exposure of staff to COVID-19 infection and to lessen use of personal protective equipment (PPE).

**Figure 2:**
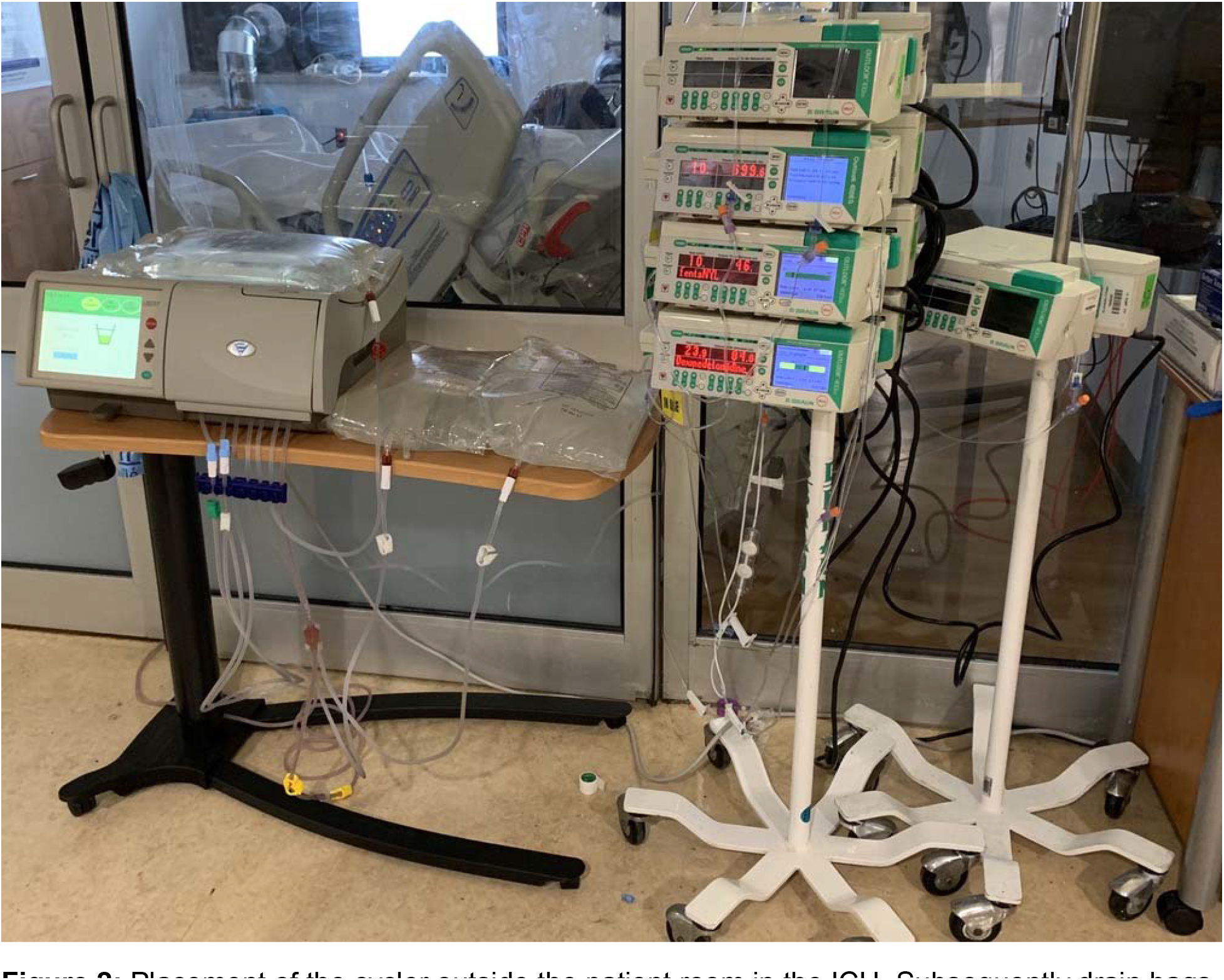
Placement of the cycler outside the patient room in the ICU. Subsequently drain bags were also used obviating the need for a drain line. The room is retrofitted with high-efficiency particulate absorbing (HEPA) filters to accommodate airborne isolation.

## Eligibility

A decision-making tree for choosing dialytic modality is shown in Figure 3. All patients who needed KRT in the ICU were eligible to receive PD catheters except for those in whom we anticipated technical challenges, usually because of prior abdominal surgery or known varices. If patients were hyperkalemic (serum potassium concentration greater than 6.5 mEq/L) despite medical therapy, such that rapid dialytic removal of potassium was necessary, CVVH treatments were started while simultaneously having a PD catheter placed if they had no contraindications. There were early concerns that respiratory status might be adversely affected by PD^17,18^. This did not occur in our patients. We were able to successfully place PD catheters in patients with morbid obesity, up to a body mass index (BMI) of 51 kg/m^2^. Some patients who had been on CVVH and had no contraindications were transitioned to PD. Additionally, because proning was not always planned, we did not consider it a contraindication.^18^

**Figure 3:**
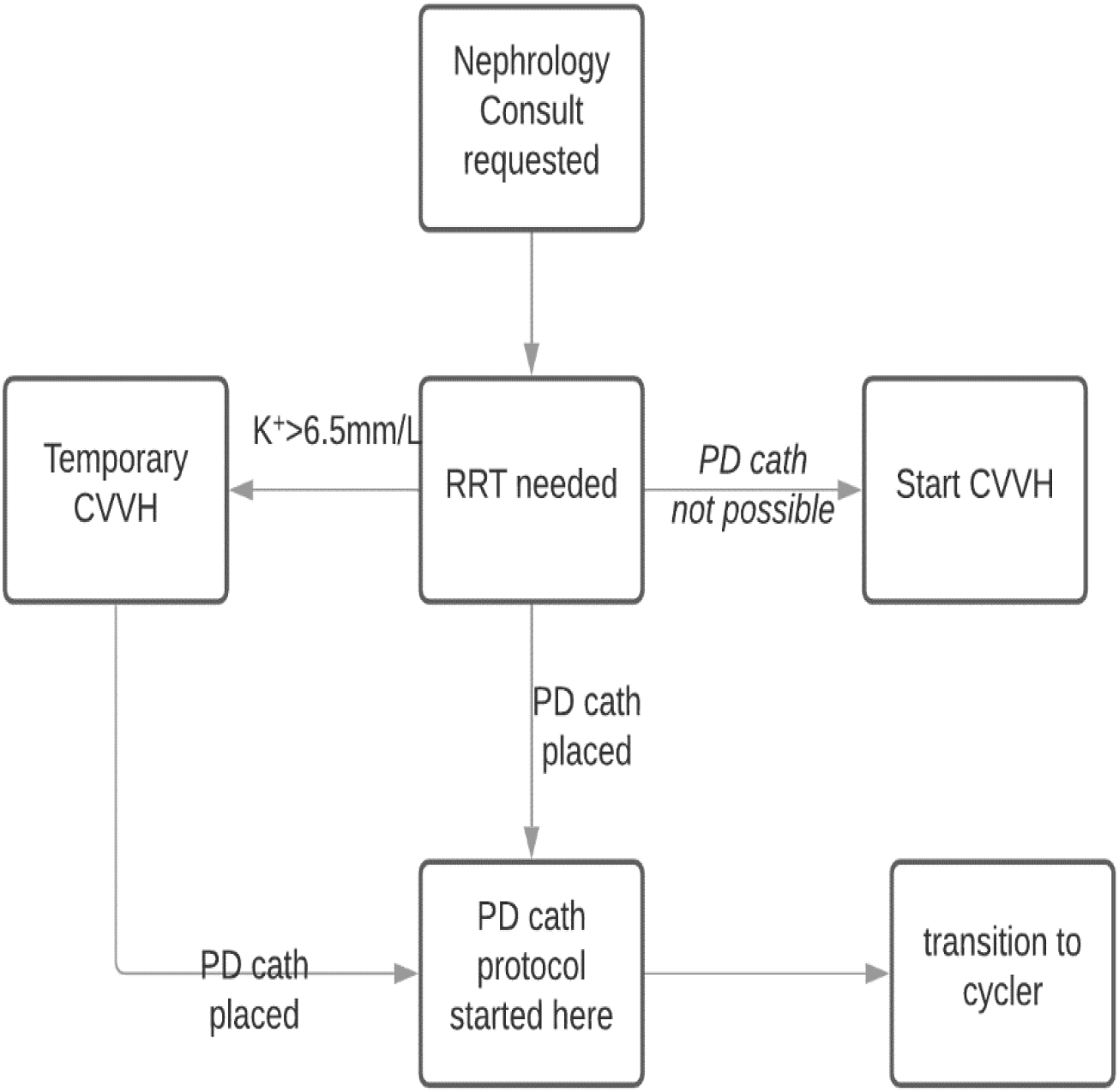
Decision-making tree for peritoneal dialysis treatment. KRT: kidney replacement therapy; CVVH: continuous venovenous hemofiltration; K: potassium.

In summary, enough PD catheters were placed to offset shortages in other modalities and allowed CVVH and hemodialysis to be done for those not suitable for PD thereby meeting the needs of all AKI patients; all patients requiring KRT received it.

## Outcomes

Daily dialysis treatments, all modalities, between April 7 – May 8, 2020 ranged from 30–40 (Figure 1). As of May 8, 2020, 63 patients were evaluated for PD and 39 PD catheters were placed into 10 women and 29 men. The average age was 59.5 years. Two patients had ESKD. Outcomes are summarized in Table 1. As of June 10, 2020, 39% of the AKI patients started on PD recovered kidney function. The average age of men and women who recovered renal function was 56 and 59.5 years, respectively, and for men and women who expired was 71.8 and 66.2 years, respectively. One ESKD patient who changed to PD because of vascular access complications was discharged on PD.

**Table 1:** Patient outcomes by gender and age. *: ESKD patients are not included in this calculation.

| **Outcome** | **Male** | **Female** | **Total** |
| --- | --- | --- | --- |
| Patient who received PD catheters | 29 | 10 | 39 |
| Recovered prior to starting PD | 1 | 0 | 1 |
| ESKD | 1 | 1 | 2 |
| Recovered and had catheter removed* | 9/27 (33%) | 5/9 (56%) | 14/36(39%) |
| Expired on PD | 20/28 (71%) | 4/10 (40%) | 24/38 (63%) |
| Average age of all patients (years) | 59.5 | 66.2 | 59.5 |
| Average age, recovered patients | 56 | 59.5 | 57.6 |
| Average age, expired patients | 71.8 | 62.3 | 60.6 |

Of the 39 patients who had catheters placed, nine had transient leaks that were resolved with reduction of dwell volume. There were no cases of peritonitis, tunnel infections or exit site infections. Two catheters needed surgical revision because of poor flow, and six catheters had minimal post placement bleeding treated with Surgicel. One patient with a poorly functioning catheter required conversion to HD before recovery.

PD delivery in our patient population was monitored closely by the attending nephrologists. The patients were monitored for extracellular fluid volume overload and depletion, electrolyte and urea levels and acid-base status to assess efficiency as with the other KRT modalities. The goal ultrafiltration (UF) volume was discussed with the ICU team and dialysate solution dextrose concentrations were adjusted accordingly. We were able to routinely achieve prescribed UF rates, removing up to 5 L in a 24-hour period, on par with other modalities. Of particular note, we used lower volume exchanges to avoid respiratory compromise.^16,17^ PD was tolerated by ventilated patients with hemodynamic instability and did not cause blood loss or systemic infections seen with the other modalities.

With this protocol and a large team of people who were able to perform many exchanges per day, we were able to maintain adequate clearance in the acute PD patients. Other than one patient switched to hemodialysis due to catheter malfunction, no PD patient required supplemental dialytic support with hemodialysis or CVVH.

## Discussion

New York City was the epicenter for COVID-19 infections in the United States in mid-March until end of May 2020. BH, the largest public hospital in New York City, and the tertiary referral hospital for the Health and Hospitals Corporation (H&H) network of New York City public hospitals was particularly taxed. The number of COVID-associated AKI patients overwhelmed our typically used dialysis modalities, compelling us to start an acute PD program to provide adequate KRT. To our knowledge, our experience with acute PD at a single hospital is the largest reported during the pandemic and one of the largest case series of acute PD reported in the United States in recent years.

Acute PD as a modality to treat KRT has become underutilized. We summarize some of the limitations to the use of various modalities in Table 2. Several studies and meta-analyses show PD to be non-inferior to IHD or CVVH.^16,17^ It also continues to be used widely in children.^19–21^ Nevertheless, there is a reluctance to use PD to treat adult patients in the ICU in the US. The reasons for this underutilization may be lack of familiarity with the technique by nephrologists, intensivists and nursing staff, and the ease of ordering CVVH by the physicians. Unease about the certainty of UF and clearance potential and misconceptions regarding complications or effectiveness despite many positive trials also contribute.^21^ We observed a mortality rate of 63% for patients with stage 3 AKI receiving PD, comparable to or less than the mortality reported in other series of COVID patients with stage 3 AKI suggesting that we were able to deliver adequate therapy.^5,22,23^ Furthermore, we were able to achieve this with a negligible complication rate, which is a tribute to the skill of the surgical team and the scrupulous technique of the PD and nursing staff.

**Table 2:** Limitations of different KRT modalities during the COVID-19 surge. ED: emergency department; HD: Hemodialysis; CVVH: continuous venovenous hemofiltration; ARDS: acute respiratory distress syndrome; ICU: intensive care units; PD: peritoneal dialysis; PA: physician assistant.

| Resource | HD | CVVH | Acute PD |
| --- | --- | --- | --- |
| Acceptance | Familiar, commonly used | Familiar, commonly used | Not used in Bellevue except rarely for chronic PD patient |
| Nursing staff | Limited trained dialysis nurses and many sick from COVID-19 | ICU nurses trained in CVVH<br>But ICU nurses overwhelmed with increased patient number | ICU nurses trained in PD, but rare use. Required retraining effort when already overwhelmed |
| Non nursing Staff | Difficult to train acutely | Difficult to train. Trained non-ICU nurses but needed significant oversight by the ICU nurses | Easier to train medical staff. Nephrologist-trained deployed MD's, PA's, and non-dialysis nurses |
| MD staffing | Adequate number of nephrologists to oversee | Adequate number of nephrologists and intensivists to oversee | Increased Nephrology staff needed to implement the program. Surgeons available due to canceled elective cases |
| Non-disposable Equipment | Fixed number of dialysis machines | Fixed number of CVVH machines | Not needed for manual PD, initially no cyclers, obtained 18 cyclers |
| Disposable equipment | Adequate supplies | Filters depleted by clotting and using machines for two people per day. Filters and fluid rationed by supplier | Percutaneous insertion kits not available. Used OR kits with open approach for placement<br>Initial supplier rationed PD supplies<br>Ordered from alternative supplier. |
| Disease related limitations | Hemodynamic instability | Hypercoagulability with significant number of clotted filters | Hypermetabolic, prone positioning was used, laparoscopic placement avoided, ARDS on ventilator |
| Benefits | Easy placement of dialysis catheter | Easy placement of dialysis catheters | PD catheter placed at the bedside. Vascular access sepsis risk avoided, less blood loss, biocompatible<br>PD skills easier to learn. Gradual volume removal |
| Potential risks | Sepsis, clotting, blood loss, hypotension, bioincompatible membrane | Sepsis, clotting, blood loss, hypocalcemia, alkalosis, potentially bioincompatible | Infection (peritonitis, tunnel infection), leak, inflow/outflow problems, hyperglycemia, bleeding at surgical site |
| Other issues | Requires plumbing in room | Not available in the ED overflow ICU due to plumbing issues | 24 hours before peritoneum primed for effective dialysis |

Our experience provides a roadmap for responses to future crises with heavy burdens of AKI. It demonstrates that the rapid development of PD capability is a viable alternative to reliance on expanding hemodialysis and CVVH capacity, and can be implemented in centers with minimal prior experience with PD. There are several advantages of this approach. We reduced our reliance on a single source of consumable supplies, a critical factor if future crises challenge typical supply chains in the same fashion as COVID-19. The simplicity of PD allows rapid training of traditional and non-traditional medical staff to deliver PD, which is not possible with more technically complex hemodialysis and CVVH options. The use of automated cyclers further simplified delivery and limited the number of patient contacts/day, thereby reducing provider risk compared to hemodialysis and CVVH and preserving PPE. Lastly, PD can be delivered manually and is not limited by the availability of dedicated machines, or electrical power.

Key elements required for successful implementation include organization of a multidisciplinary team including nephrology, surgical and nursing, development of standard protocols, education of ICU staff and a resource for rapid training. We believe that adoption of the steps outlined may be key to avoiding the need to ration KRT in future waves of COVID-19 or other health crises and should be considered for programs considering how to ensure adequate responses. We advocate that acute PD can and should be used in acutely ill patients. During times of shortages, it can be used to offset other modalities when expanding current resources is impossible.

Our experience demonstrates that establishing an acute PD program during a crisis is possible and can be lifesaving.

## Data Availability

none

## Acknowledgements

We thank all of the ‘PD team members’, especially Joyce Kadji (one of the founding members) for all their hard work and dedication during this difficult time. We are grateful for the support of the Bellevue administration. Thank you for Fresenius for supplying PD supplies during the crisis. Thank you, Dr. K.H. Gardner, for assistance with editing and formatting. Thanks to J. Uribarri for all his ongoing advice and support.

## Key Points

### Question

Can acute peritoneal dialysis (PD) be used to complement traditional kidney replacement therapy (KRT) modalities during a crisis situation?

### Findings

In the midst of the 2020 COVID-19 pandemic, we quickly assembled an acute PD response team and started 38 patients on treatment despite lack of prior routine use. As of June 10, 2020, 16 patients recovered renal function and no major complications were found.

### Meaning

Despite severe shortages of staff, supplies and dialysis machines during the COVID-19 pandemic, peritoneal dialysis provided adequate kidney replacement therapy to all admitted patients who needed it and allowed us to avoid rationing of care.

